# Dementia and End-of-Life Shared Decision-Making Among Older US Adults

**DOI:** 10.64898/2026.03.27.26349555

**Authors:** Zhigang Xie, Young-Rock Hong, Melissa J. Armstrong, Xiangren Wang, Molly Jacobs

## Abstract

**Introduction:** End-of-life decision-making poses unique challenges for individuals with dementia and their family caregivers as cognitive decline shifts decision-making responsibility to surrogates.

**Methods:** Using 2010–2022 Health and Retirement Study (HRS) exit interview data, we compared advance directive completion, decision-making needs near death, involvement of others in decision-making, and concordance between expressed preferences and care received among decedents with and without dementia. Analyses incorporated HRS exit interview sampling weights, primary sampling units, and strata to account for the complex multistage probability design of HRS and produce nationally representative estimates of U.S. older adult decedents (≥50 years). Weighted descriptive statistics and design-adjusted Wald tests were used to compare groups.

**Results:** Among 5,389 decedents, 1,010 (weighted 17.7%) had dementia prior to death. Decedents with dementia were more likely to have completed advance directives than those without dementia (81.3% vs. 69.1%, p<.001). However, they also had significantly higher decision-making needs in the final days of life (54.3% vs. 47.2%, p<.001). Children or grandchildren were more frequently involved in care decisions for decedents with dementia (63.9% vs. 45.6%, p<.001). Despite differences in decision-making processes, most decedents in both groups expressed preferences for comfort-focused care, and preference–care concordance exceeded 90% in both groups.

**Conclusions:** Findings suggested that dementia reshaped the structure and intensity of the shared decision-making process by increasing surrogate engagement and decisional demands, underscoring the importance of early advance care planning and structured support for family caregivers to sustain goal-concordant care.

## Introduction

Shared decision-making is increasingly recognized as a central mechanism for delivering high-quality care across the healthcare continuum by fostering awareness of available options, supporting deliberation about care alternatives, and ensuring that clinical decisions reflect patients’ values, goals, and informed preferences.^1-3^ In the context of serious illness and end-of-life care, there have been growing calls to improve the quality of decision-making for patients who lose decisional capacity, including through advance care planning discussions, completion of advance directives, and greater engagement of family caregivers as surrogates.^4-6^ Despite these efforts, substantial challenges in end-of-life shared decision-making remain well documented. Studies have reported low rates of advance care planning discussions,^7,8^ inadequate communication between physicians and surrogate decision makers,^9^ conflicts between patient values and medical recommendations, and disagreements between clinicians and family members regarding the care of patients who ultimately die.^10^

Dementia presents a particularly complex context for end-of-life shared decision-making.^10^ As the disease progresses, cognitive decline gradually impairs patients’ ability to participate in complex medical decisions, shifting responsibility for decision-making to family caregivers or other surrogates.^11^ At the same time, individuals with dementia often experience prolonged illness trajectories characterized by uncertainty, progressive functional decline, and multiple care transitions, which frequently require decisions about life-sustaining treatments, hospitalization, feeding interventions, and hospice care.^12-14^ These decisions are ideally guided by patients’ previously expressed values and preferences. However, in practice, surrogates often face substantial challenges interpreting patients’ wishes while navigating emotional stress, incomplete information, and varying levels of clinician communication and support.^15^ As a result, end-of-life decision-making for people with dementia frequently occurs under conditions of uncertainty and may not always reflect patients’ goals of care.^14-16^

Despite growing recognition of the importance of shared decision-making in serious illness, most existing shared decision-making frameworks have not been developed specifically for the context of dementia.^17^ Consequently, how to effectively support people living with dementia and their caregivers in shared decision-making remains unclear. ^18,19^ Existing studies have largely focused on decision-making within dementia populations or specific care settings (e.g., nursing homes or hospitals), rather than comparing end-of-life decision-making processes between individuals with and without dementia at a population level.^20-25^ Understanding how dementia shapes end-of-life decision-making relative to individuals without dementia is critical for informing policies and clinical practices that address the growing population of people living with dementia.

This study uses nationally representative data from the Health and Retirement Study (HRS) exit interviews from 2010–2022 to compare key aspects of end-of-life decision-making among decedents with and without dementia. Specifically, we examine differences in advance directive completion, decision-making needs in the final days of life, involvement of others (e.g., spouses, children, etc.) in decision-making, and concordance between stated end-of-life care preferences and the care received.

## Methods

### Data Source and Study Population

We used data from the HRS exit interviews collected between 2010 and 2022.^26,27^ HRS is a nationally representative longitudinal survey of adults in the United States (U.S.) aged 50 years and older that employs a multistage area probability sampling design.^28,29^ When a participant dies, a proxy respondent (typically a family member or close caregiver) completes an exit interview describing the decedent’s health, end-of-life care experiences, and decision-making processes near death.^28,29^ During the study period, there were 9,518 decedents. The analytic sample included respondents with complete information on dementia status, end-of-life decisional needs, advance directives, and selected decedent characteristics (Table 1). After applying these criteria, the sample was reduced to 6,398 decedents. We then excluded an additional 1,009 individuals who did not have positive sampling weights, resulting in a final analytic sample of 5,389 decedents (representing 22,231,275 weighted decedents) (Figure 1). The University of Florida Institutional Review Board determined that this study involved non-human subjects research using publicly available, de-identified data and was therefore exempt from review.

**Figure 1:**
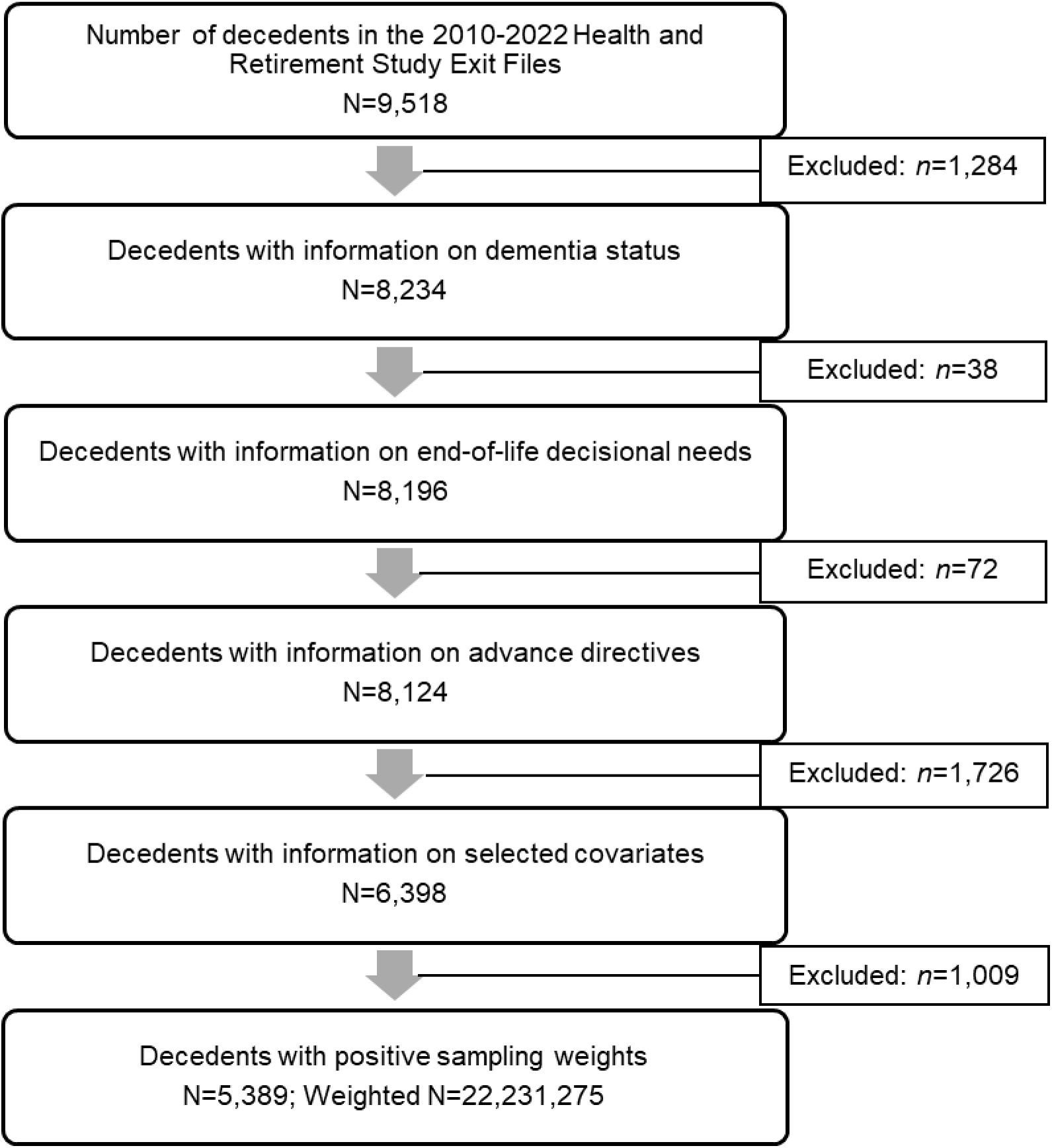
Study Sample Selection Process.

**Table 1.** Respondent-Reported Characteristics of Decedents by Dementia Status Before Death (n = 5,389, Weighted n = 22,231,275)

| Characteristics | Dementia (Yes) |  | Dementia (No) |  | *p* |
| --- | --- | --- | --- | --- | --- |
|  | No. | Weighted % | No. | Weighted % |  |
| **Study Sample** |  |  |  |  |  |
| Unweighted, n | 1010 |  | 4379 |  |  |
| Weighted, n (%) | 3,933,814 | 17.7 | 18,297,461 | 82.3 |  |
| **Age of Death (Year)** |  |  |  |  | <.001 |
| 50-64 | 52 | 6.0 | 616 | 17.1 |  |
| 65-84 | 463 | 44.6 | 2320 | 50.9 |  |
| 85+ | 495 | 49.4 | 1443 | 32.0 |  |
| **Sex** |  |  |  |  | <.001 |
| Male | 425 | 42.5 | 2188 | 51.3 |  |
| Female | 585 | 57.5 | 2191 | 48.7 |  |
| **Race/ethnicity** |  |  |  |  | .005 |
| NH White | 700 | 77.2 | 3167 | 80.2 |  |
| NH Black | 205 | 14.2 | 748 | 10.4 |  |
| Hispanic | 91 | 6.9 | 366 | 6.5 |  |
| Other | 14 | 1.7 | 98 | 2.9 |  |
| **Education** |  |  |  |  | .026 |
| GED or Lower | 384 | 32.8 | 1451 | 29.8 |  |
| High School Graduate | 290 | 28.8 | 1343 | 30.7 |  |
| Some Colleges | 184 | 21.2 | 925 | 22.2 |  |
| College and above | 152 | 17.0 | 659 | 17.2 |  |
| **Marital Status** |  |  |  |  | <.001 |
| Married or Living with a Partner | 420 | 41.1 | 2217 | 50.7 |  |
| Divorced or Separated | 105 | 11.8 | 496 | 12.5 |  |
| Widowed | 456 | 43.0 | 1479 | 30.8 |  |
| Never Married | 29 | 4.1 | 187 | 6.0 |  |
| **Income (Percentile)** |  |  |  |  | .118 |
| 75.0%-100% | 252 | 26.4 | 1165 | 28.4 |  |
| 50-74.9% | 55 | 5.2 | 269 | 6.5 |  |
| 25-49.9% | 474 | 47.9 | 1906 | 42.9 |  |
| <25% (lowest income) | 229 | 20.5 | 1039 | 22.2 |  |
| **Death Cause** |  |  |  |  | <.001 |
| Cancer | 134 | 13.6 | 1282 | 29.9 |  |
| Cardiovascular Disease | 340 | 32.8 | 1517 | 34.4 |  |
| Other | 536 | 53.6 | 1580 | 35.8 |  |
| **Death Expected** |  |  |  |  | <.001 |
| Yes | 707 | 70.6 | 2450 | 56.3 |  |
| No | 303 | 29.4 | 1929 | 43.7 |  |

Abbreviation: NH, non-Hispanic

### Measures

Dementia status was classified as yes or no based on proxy reports indicating that the decedent had Alzheimer’s disease or another form of dementia before death. The two primary binary outcomes were advance directive completion and decision-making needs near death. Advance directive completion was defined as whether the decedent had written instructions regarding medical treatment preferences or appointed a surrogate decision maker, including documents such as a living will or durable power of attorney for healthcare. Decision-making needs were measured based on the question of whether important medical decisions needed to be made during the final days of life.

We also examined several secondary outcomes including: (1) whether the decedent had the capacity to be involved in medical decision-making near the end of life (yes/no); (2) the primary decision maker responsible for end-of-life medical decisions (decedent, spouse/partner, child or grandchild, or others—including other family members, friends, or clinicians); and (3) concordance between preferred and received comfort-focused care in the final days of life, measured by comparing proxy reports of the decedent’s stated preference for comfort-focused care with the care actually received.

Demographic characteristics included age at death (50–64, 65–84, ≥85 years), sex (male, female), race/ethnicity (non-Hispanic White, non-Hispanic Black, Hispanic, non-Hispanic Other), educational attainment (GED or lower, high school graduate, some college, college graduate or above) marital status (married or living with a partner, divorced or separated, widowed, never married), income level (with lower percentiles indicating lower income), cause of death (cancer, cardiovascular disease, or other causes), and whether the death was expected (yes/no). These variables were selected based on prior literature demonstrating their associations with advance care planning and end-of-life decision-making.^7,9^

### Statistical Analysis

All analyses incorporated HRS exit interview sampling weights from the decedent’s last interview wave, along with survey strata and primary sampling units, to account for the complex multistage sampling design and to produce nationally representative estimates. All statistical analyses were conducted using SAS software (version 9.4; SAS Institute Inc) and Stata (version 19; StataCorp). Two-sided p-values <0.05 were considered statistically significant.

First, we calculated survey-weighted descriptive statistics to compare demographic and clinical characteristics of decedents with and without dementia. Second, we estimated survey-weighted prevalence for key end-of-life decision-making measures, including decision-making needs near death, advance directive completion, decedents’ capacity to participate in end-of-life medical decisions, the corresponding primary decision maker, and concordance between preferred and received palliative or comfort-focused care in the final days of life. Finally, we used survey-weighted modified Poisson regression models with robust variance estimation to estimate adjusted risk ratios (aRRs) and 95% confidence intervals for factors associated with (1) decision-making needs near death and (2) advance directive completion. Multivariable models adjusted for sociodemographic characteristics and dementia status (listed in Table 1).

## Results

### Sample Characteristics

Among 5,389 decedents, 1,010 (17.7%) had dementia prior to death, representing approximately 3.9 million U.S. decedents, while 4,379 (82.3%) did not have dementia (Table 1). Decedents with dementia were significantly older at death compared with those without dementia (p<.001). Nearly half of individuals with dementia died at age 85 years or older (49.4%), compared with 32.0% among those without dementia. Women were more likely to die with dementia than men (57.5% vs. 48.7%, p<.001). In addition, a significantly higher proportion of decedents with dementia were non-Hispanic Black (14.2% vs. 10.4%, p=.005), had a GED or lower level of education (32.8% vs. 29.8%, p=.026), and were widowed at the time of death (43.0% vs. 30.8%, p=.005). Compared with decedents without dementia, those with dementia were also more likely to have causes of death other than cancer or cardiovascular disease (53.6% vs. 35.8%, p<.001), and to have deaths that were expected (70.6% vs. 56.3%, p<.001).

### Advance Directive Completion

Advance directive completion was common in the study sample but differed substantially by dementia status (Table 2). Individuals with dementia had a significantly higher prevalence of advance directives compared with those without dementia (81.3% vs. 69.1%, p<.001). After adjustment for demographic characteristics, dementia remained associated with a higher likelihood of advance directive completion (aRR=1.11, 95% CI: 1.07–1.15). Advance directive completion also increased markedly with age. Compared with decedents aged 50–64 years, those aged 65–84 years were 28% more likely to have completed an advance directive (aRR=1.28, 95% CI: 1.16–1.41), and those aged 85 years or older were 49% more likely to have done so (aRR=1.49, 95% CI: 1.34–1.64). Significant racial and ethnic disparities were also observed. Non-Hispanic Black (aRR=0.70, 95% CI: 0.64–0.77) and Hispanic (aRR=0.73, 95% CI: 0.65–0.82) decedents were significantly less likely to have completed advance directives compared with non-Hispanic White decedents. Additionally, decedents whose deaths were unexpected were less likely to have completed advance directives compared with those whose deaths were expected (aRR=0.85, 95% CI: 0.81–0.89).

**Table 2.**
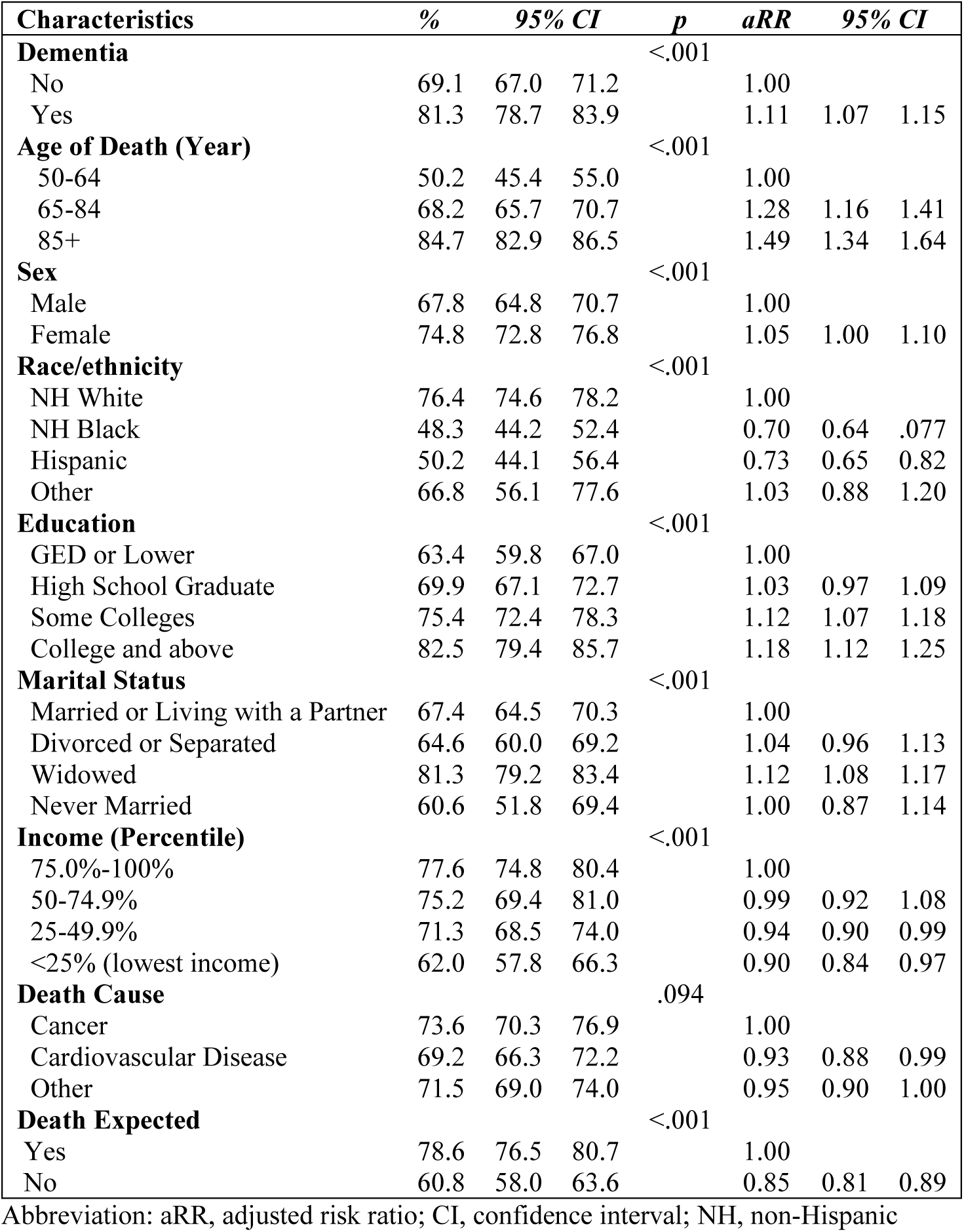
Prevalence and Determinants of Advance Directives Among Decedents Using Survey-Weighted Modified Poisson Regression (n = 5,389, Weighted n = 22,231,275)

### Decision-Making Needs and Surrogate Involvement

Overall, a higher proportion of decedents with dementia lacked the capacity to participate in medical decision-making during the final days of life compared with those without dementia (78.9% vs. 48.0%, p<.001) (Figure 2). Correspondingly, children or grandchildren were more frequently reported as the primary decision makers for decedents with dementia (63.9% vs. 45.6%, p<.001). Decision-making needs in the final days of life were common among decedents. The prevalence of decision-making needs was significantly higher among individuals with dementia compared with those without dementia (54.3% vs. 47.2%, p<.001) (Table 3). In adjusted analyses, dementia remained significantly associated with a higher likelihood of decision-making needs (aRR=1.09, 95% CI: 1.01–1.19). Educational attainment was also associated with decision-making needs. Compared with decedents with a GED or lower level of education, those with a college degree or higher had a greater likelihood of experiencing decision-making needs near death (aRR=1.19, 95% CI: 1.09–1.31). In contrast, decedents whose deaths were unexpected were less likely to have decision-making needs compared with those whose deaths were expected (aRR=0.80, 95% CI: 0.75–0.86).

**Figure 2.**
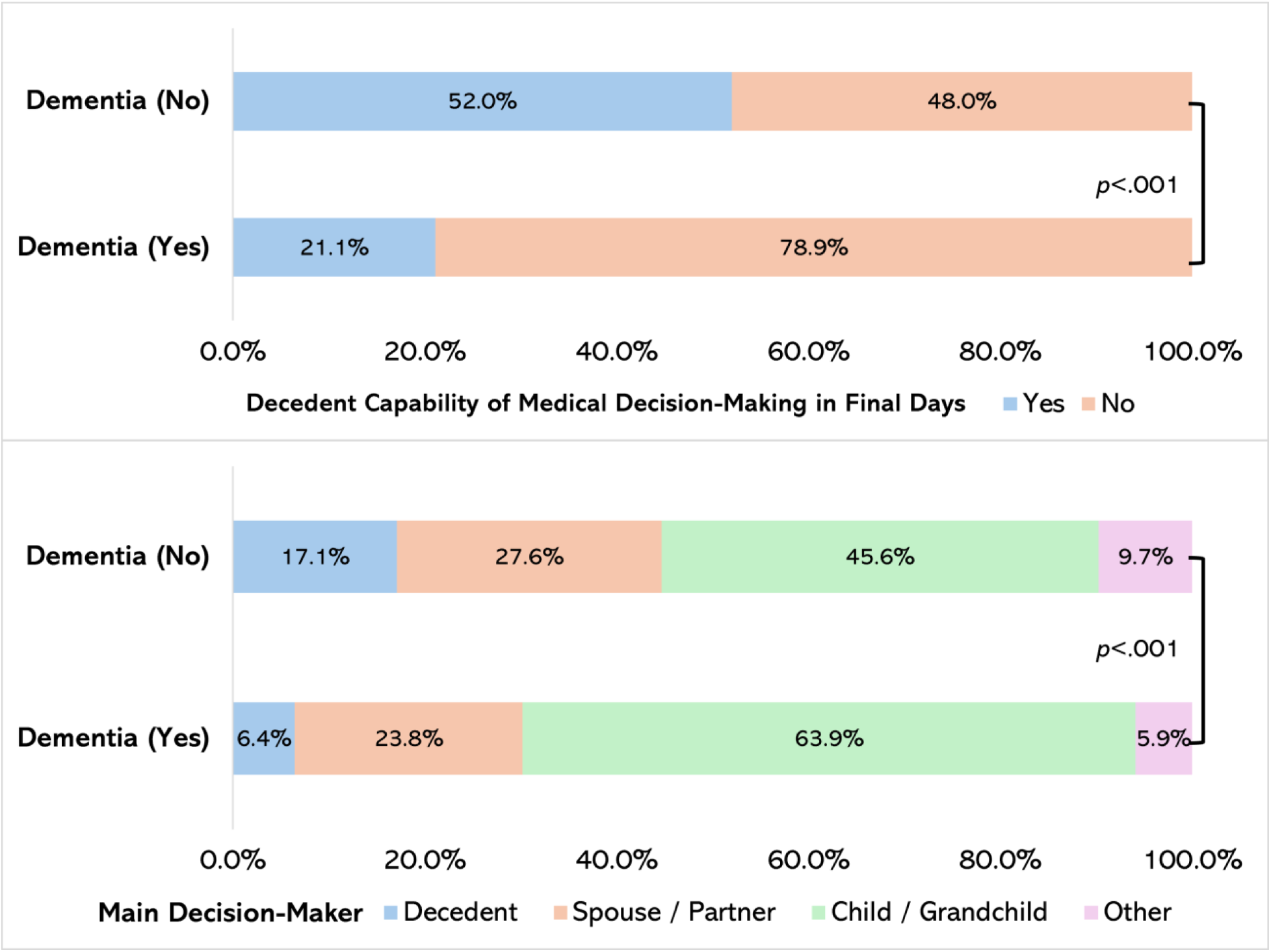
Decedents’ Capability for Involvement in End-Of-Life Medical Care Decisions and the Corresponding Primary Decision-Maker (n = 1,451, Weighted n = 5,829,094)

**Table 3.** Prevalence and Determinants of Decisional Needs Among Decedents Using Survey-Weighted Modified Poisson Regression (n = 5,389, Weighted n = 22,231,275)

| **Characteristics** | **Prevalence %** | **95% CI** |  | **p** | **aRR** | **95% CI** |  |
| --- | --- | --- | --- | --- | --- | --- | --- |
| **Dementia** |  |  |  | <.001 |  |  |  |
| No | 47.2 | 45.6 | 48.8 |  | 1.00 |  |  |
| Yes | 54.3 | 50.9 | 57.7 |  | 1.09 | 1.01 | 1.19 |
| **Age of Death (Year)** |  |  |  | .017 |  |  |  |
| 50-64 | 45.6 | 40.8 | 50.4 |  | 1.00 |  |  |
| 65-84 | 46.9 | 44.6 | 49.3 |  | 0.99 | 0.87 | 1.13 |
| 85+ | 51.8 | 49.7 | 54.0 |  | 1.04 | 0.89 | 1.21 |
| **Sex** |  |  |  | <.001 |  |  |  |
| Male | 45.7 | 43.6 | 47.8 |  | 1.00 |  |  |
| Female | 51.2 | 49.2 | 53.1 |  | 1.11 | 1.03 | 1.19 |
| **Race/ethnicity** |  |  |  | .005 |  |  |  |
| NH White | 49.6 | 48.0 | 51.1 |  | 1.00 |  |  |
| NH Black | 41.2 | 36.8 | 45.7 |  | 0.89 | 0.96 | 1.15 |
| Hispanic | 45.6 | 40.4 | 50.8 |  | 0.99 | 0.87 | 1.13 |
| Other | 51.5 | 41.2 | 61.9 |  | 1.10 | 0.90 | 1.35 |
| **Education** |  |  |  | <.001 |  |  |  |
| GED or Lower | 44.4 | 42.0 | 46.7 |  | 1.00 |  |  |
| High School Graduate | 47.7 | 45.1 | 50.3 |  | 1.05 | 0.96 | 1.15 |
| Some Colleges | 49.8 | 46.4 | 53.2 |  | 1.09 | 0.99 | 1.21 |
| College and above | 55.1 | 51.2 | 58.9 |  | 1.19 | 1.09 | 1.31 |
| **Marital Status** |  |  |  | .140 |  |  |  |
| Married or Living with a Partner | 47.1 | 44.9 | 49.3 |  | 1.00 |  |  |
| Divorced or Separated | 50.9 | 45.2 | 56.6 |  | 1.11 | 0.98 | 1.27 |
| Widowed | 50.4 | 48.0 | 52.9 |  | 1.03 | 0.95 | 1.12 |
| Never Married | 43.3 | 35.8 | 50.8 |  | 0.94 | 0.79 | 1.11 |
| **Income (Percentile)** |  |  |  | .055 |  |  |  |
| 75.0%-100% | 51.3 | 48.0 | 54.5 |  | 1.00 |  |  |
| 50-74.9% | 50.3 | 43.1 | 57.5 |  | 1.00 | 0.85 | 1.19 |
| 25-49.9% | 48.4 | 46.0 | 50.8 |  | 0.97 | 0.88 | 1.06 |
| <25% (lowest income) | 44.3 | 40.9 | 47.8 |  | 0.91 | 0.83 | 1.00 |
| **Death Cause** |  |  |  | .011 |  |  |  |
| Cancer | 50.0 | 46.7 | 53.2 |  | 1.00 |  |  |
| Cardiovascular Disease | 44.6 | 41.3 | 47.8 |  | 0.95 | 0.85 | 1.06 |
| Other | 50.8 | 48.4 | 53.1 |  | 1.04 | 0.95 | 1.15 |
| **Death Expected** |  |  |  |  |  |  |  |
| Yes | 53.5 | 51.8 | 55.3 |  | 1.00 |  |  |
| No | 41.1 | 38.8 | 43.5 |  | 0.80 | 0.75 | 0.86 |

Abbreviation: aRR, adjusted risk ratio; CI, confidence interval; NH, non-Hispanic

### Preference-Care Concordance

Finally, among decedents who had completed advance directives, both those with and without dementia reported high levels of preference for palliative care in the final days of life (90.6% vs. 89.4%, p=.432) and high levels of receipt of palliative care (94.6% vs. 93.5%, p=.411), with no statistically significant differences observed between the groups (Figure 3).

**Figure 3.**
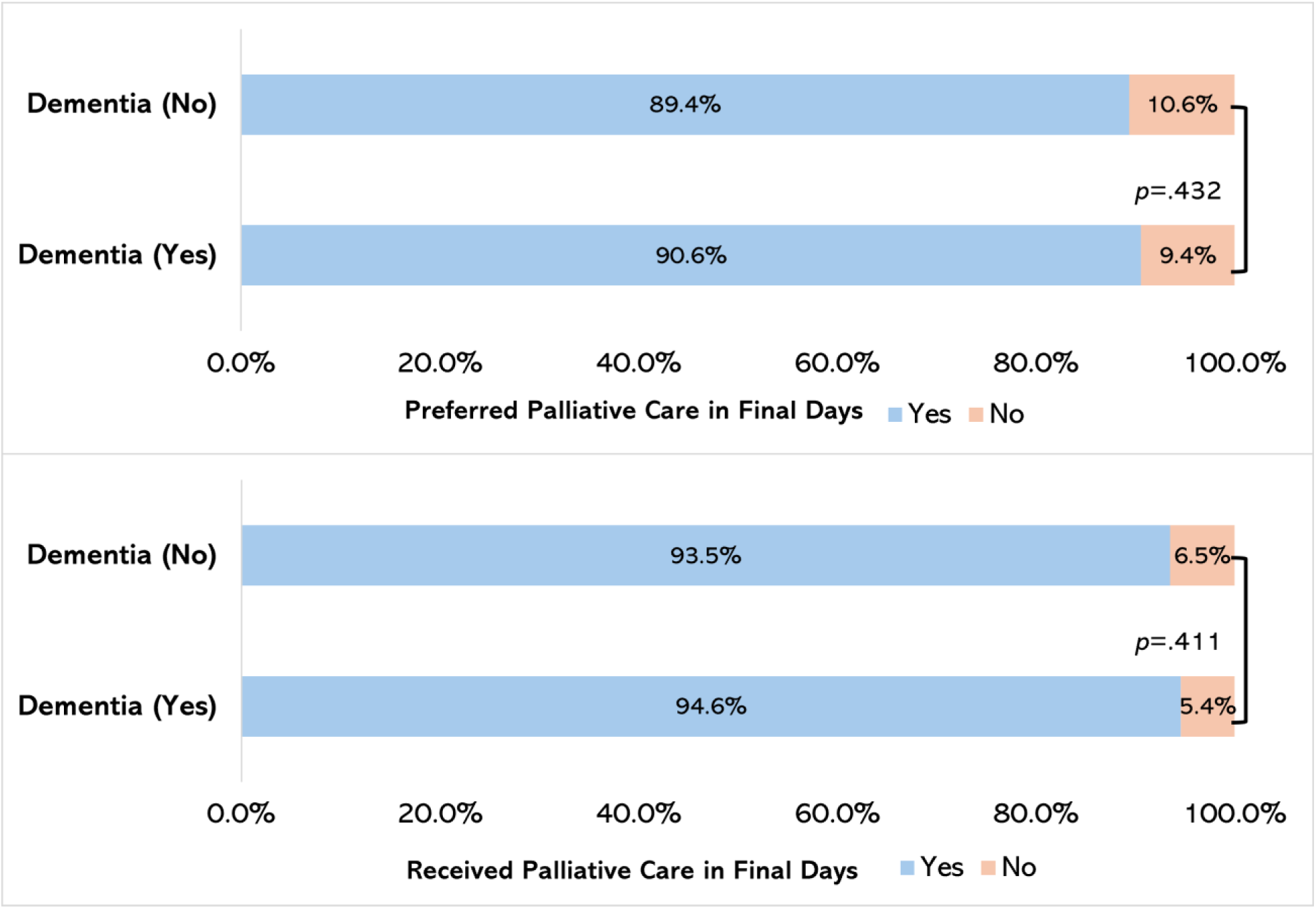
Prevalence of Decedents’ Preferred Versus Received Palliative Care During the Final Days of Life (n = 749, Weighted n = 3,078,579)

## Discussion

In this nationally representative study of older U.S. decedents, dementia was associated with a distinct end-of-life shared decision-making pattern characterized by greater advance care planning, increased decisional burden, and greater reliance on surrogate decision makers. Although individuals with dementia were more likely to have completed advance directives, they also experienced higher levels of decision-making needs near death and were substantially less likely to retain decisional capacity. These findings highlight a critical paradox in end-of-life care for dementia: formal documentation of preferences does not fully mitigate the complexity or intensity of real-time decision-making.

From a mechanistic perspective, dementia fundamentally alters the shared decision-making process through progressive cognitive decline, which shifts decision authority from patients to surrogates.^30-32^ Even when advance directives are present, they may lack the specificity required to guide decisions about acute clinical events, such as infections, hospitalizations, or feeding interventions, which are common in advanced dementia.^13,33,34^ As a result, surrogate decision makers (most often children or grandchildren in this study) must interpret prior wishes and apply them to evolving clinical contexts. This process likely contributes to the higher observed prevalence of decision-making needs among decedents with dementia, despite higher rates of documented preparation.^35,36^

Importantly, prior conceptualizations of shared decision-making as an iterative process requiring continuous communication, deliberation, and reassessment of patient values and goals.^37-39^ In serious illness contexts, particularly dementia, this process can be further complicated by progressive loss of decisional capacity and the need for surrogate engagement across multiple care transitions.^21,40,41^ In this context, advance care planning should be further emphasized not as a one-time documentation exercise, but as a continuous process of values clarification and decision support that adapts to disease progression.^5,6^ Future studies should further investigate how shared decision-making in dementia unfolds over the disease trajectory among patient with dementia, family caregiver and clinicians.

Despite these challenges, we observed consistently high levels of concordance between expressed preferences for comfort-focused care and the care received, regardless of dementia status, which is consistent with a prior study using electronic health record sources.^42^ This is a notable and encouraging finding, suggesting that, at a population level, care systems may be reasonably effective at honoring general end-of-life preferences. However, high concordance should be interpreted cautiously. Prior work has demonstrated that agreement between documented preferences and care received may reflect broad alignment (e.g., comfort-focused vs. life-prolonging care) rather than precise concordance at the level of specific clinical decisions.^43,44^ Thus, while concordance appears high, it may mask important variability in how preferences are operationalized in practice.

### Implications

These findings have several implications for palliative care delivery and policy. First, they underscore the value of engaging family caregivers throughout the decision-making process. Given the central role of surrogates, interventions should focus on preparing patient-caregiver dyads for their roles, including improving communication skills, clarifying patients’ values, and providing decision-support tools tailored to dementia care.^15,20^ Second, healthcare systems should prioritize integration of palliative care principles earlier in the dementia trajectory, rather than reserving such support for the final stages of life.^12,45^ Early palliative care involvement may help reduce decisional burden, align care with patient goals, and support caregivers navigating complex decisions.

Third, the observed lower engagement with advance care planning among non-Hispanic Black and Hispanic populations highlight persistent inequities in access to and engagement with advance care planning.^46-48^ Prior research shows that these disparities are shaped largely by cultural beliefs, preferences for family-centered decision-making, and levels of trust in the healthcare system.^49^ For example, differences between African American and White individuals have been largely attributed to the higher levels of mistrust and experiences of discrimination reported by African Americans.^50^ Addressing these disparities will require culturally tailored interventions, community-based engagement, and efforts to build trust in healthcare systems among historically underserved populations.

Taken together, this study contributes to a growing body of literature emphasizing that achieving goal-concordant care in dementia requires not only documentation of preferences but also robust systems to support surrogate decision-making under conditions of uncertainty.^17-19^ Interventions that integrate advance care planning with caregiver support, clinician communication training, and early palliative care involvement may be particularly effective in improving the quality of end-of-life care for individuals living with dementia.

### Limitations

This study has several limitations that should be considered when interpreting the findings. First, the use of proxy-reported data from HRS exit interviews introduces the potential for recall, subjectivity, recency, and favorability bias and measurement error, particularly for subjective constructs such as decision-making needs, decisional capacity, and concordance between preferences and care received; proxy respondents, although typically close caregivers, may not fully capture patients’ perspectives or the complexity of clinical decision-making processes. Second, dementia status was based on proxy reports rather than clinical adjudication, raising the possibility of misclassification, particularly among individuals with mild or undiagnosed cognitive impairment. Third, measures of advance care planning and preference–care concordance were relatively broad and may not reflect the specificity, timing, or quality of advance directives or the nuanced alignment between stated preferences and specific treatment decisions; consequently, concordance may be overestimated if based on general goals of care rather than discrete clinical choices. Fourth, although we adjusted for a range of demographic and clinical characteristics, residual confounding may persist due to unmeasured factors such as caregiver characteristics (e.g., health literacy, burden), health system features, regional practice patterns, and the quality of clinician–family communication, all of which are known to influence shared decision-making. Fifth, the cross-sectional nature of exit interview data limits our ability to assess the temporal sequencing and evolution of advance care planning, decisional needs, and care delivery over the disease trajectory, precluding causal inference. Sixth, due to missing data (e.g., partial interviews) and the absence of valid sampling weights, a portion of the original study sample was excluded from the final analytic sample. Although initial missingness analyses indicated no significant outcome differences between included and excluded decedents, it is possible that those excluded differed systematically on other unmeasured characteristics. Finally, because HRS exit interviews rely on decedent samples, findings may not fully generalize to individuals currently living with dementia or to care experiences outside the U.S. context. Despite these limitations, the use of a large, nationally representative dataset provides important population-level insights into end-of-life decision-making among individuals with and without dementia.

## Conclusions

Dementia reshaped the structure and intensity of the shared decision-making process by increasing surrogate engagement and decisional demands, underscoring the importance of early advance care planning and structured support for family caregivers to sustain goal-concordant care. Together, these findings underscore the urgent need for healthcare systems to better equip patients with dementia, their caregivers, and clinicians with the tools required to navigate complex end-of-life decisions.

## Acknowledgements

None.

## Author Contributions

**ZX:** Conceptualization; methodology; software; formal analysis; original draft; writing – review and editing.

**YH:** Conceptualization; methodology; supervising; writing – review and editing; senior author.

**MJA:** Conceptualization; writing – review and editing.

**XW:** Conceptualization; writing – review and editing.

**MJ:** Conceptualization; original draft; supervising; writing – review and editing; corresponding senior author.

## Ethical Considerations

As the HRS is a publicly available, de-identified dataset, the Institutional Review Board (IRB) at the first author’s university determined that this study was non-human subject research.

## Consent to Participate

It is not applicable as this is a secondary data analysis with publicly available deidentified data.

## Consent for Publication

Not applicable.

## Data Availability Statement

The data supporting the findings of this study are openly available on the Health and Retirement Study website. These data were derived from the following resources available in the public domain: https://hrs.isr.umich.edu/data-products.

## References

1. Barry MJ, Edgman-Levitan S. Shared decision making—the pinnacle of patient-centered care. N Engl J Med. 2012;366(9):780–781.

2. Bravo P, Härter M, McCaffery K, Giguère A, Hahlweg P, Elwyn G. 20 years after the start of international Shared Decision-Making activities: Is it time to celebrate? Probably…. Zeitschrift für Evidenz, Fortbildung und Qualität im Gesundheitswesen. 2022;171:1–4.

3. Bomhof-Roordink H, Gärtner FR, Stiggelbout AM, Pieterse AH. Key components of shared decision making models: a systematic review. BMJ open. 2019;9(12):e031763.

4. Centers for Medicare & Medicaid Services (CMS), HHS. Medicare program; revisions to payment policies under the physician fee schedule and other revisions to part B for CY 2018; Medicare shared savings program requirements; and Medicare diabetes prevention program. Final rule. Fed Regist. 2017;82(219):52976–53371.

5. Sudore RL, Lum HD, You JJ, et al. Defining advance care planning for adults: a consensus definition from a multidisciplinary Delphi panel. J Pain Symptom Manage. 2017;53(5):821–832. e1.

6. Committee on Approaching Death, Addressing Key End-of-Life Issues. Dying in America: Improving quality and honoring individual preferences near the end of life. 2015.

7. Palmer MK, Jacobson M, Enguidanos S. Advance Care Planning For Medicare Beneficiaries Increased Substantially, But Prevalence Remained Low: Study examines Medicare outpatient advance care planning claims and prevalence. Health Aff. 2021;40(4):613–621.

8. Gotanda H, Walling AM, Zhang JJ, Xu H, Tsugawa Y. Timing and setting of billed advance care planning among Medicare decedents in 2017–2019. J Am Geriatr Soc. 2023;71(10):3237–3243.

9. Virdun C, Luckett T, Davidson PM, Phillips J. Dying in the hospital setting: a systematic review of quantitative studies identifying the elements of end-of-life care that patients and their families rank as being most important. Palliat Med. 2015;29(9):774–796.

10. Groen-van de Ven L, Smits C, Span M, et al. The challenges of shared decision making in dementia care networks. International Psychogeriatrics. 2018;30(6):843–857.

11. Miller LM, Whitlatch CJ, Lyons KS. Shared decision-making in dementia: A review of patient and family carer involvement. Dementia. 2016;15(5):1141–1157.

12. Bartley MM, Suarez L, Shafi RM, Baruth JM, Benarroch AJ, Lapid MI. Dementia care at end of life: current approaches. Curr Psychiatry Rep. 2018;20(7):50.

13. Gozalo P, Teno JM, Mitchell SL, et al. End-of-life transitions among nursing home residents with cognitive issues. N Engl J Med. 2011;365(13):1212–1221.

14. Triandafilidis Z, Carr S, Davis D, et al. What care do people with dementia receive at the end of life? Lessons from a retrospective clinical audit of deaths in hospital and other settings. BMC geriatrics. 2024;24(1):40.

15. Fetherstonhaugh D, McAuliffe L, Bauer M, Shanley C. Decision-making on behalf of people living with dementia: how do surrogate decision-makers decide? J Med Ethics. 2017;43(1):35–40.

16. Geerse OP, Lamas DJ, Bernacki RE, et al. Adherence and concordance between serious illness care planning conversations and oncology clinician documentation among patients with advanced cancer. J Palliat Med. 2021;24(1):53–62.

17. Peisah C, Sorinmade OA, Mitchell L, Hertogh CM. Decisional capacity: toward an inclusionary approach. International Psychogeriatrics. 2013;25(10):1571–1579.

18. Daly RL, Bunn F, Goodman C. Shared decision-making for people living with dementia in extended care settings: a systematic review. BMJ open. 2018;8(6):e018977.

19. Mattos MK, Gibson JS, Wilson D, Jepson L, Ahn S, Williams IC. Shared decision-making in persons living with dementia: A scoping review. Dementia. 2023;22(4):875–909.

20. Porteny T, Lynch M, Covaleski A, et al. Medical decision-making experiences of persons with dementia and their carepartners: a qualitative study. BMC Palliative Care. 2025;24(1):99.

21. Kim H, Cho J, Park J, Kim SS. Surrogate Decision-Making Practices Regarding End-of-Life Care for People With Dementia in Long-Term Care Hospitals: A Qualitative Descriptive Study. J Adv Nurs. 2025.

22. Scheeres-Feitsma TM, Schaafsma P, van Kempen J, van der Steen JT. Bringing up the end of life and euthanasia. A mixed method study on consultations with people with dementia and their families in the hospital setting. Palliat Med. 2026:02692163261416275.

23. Cincidda C, Pizzoli SFM, Ongaro G, Oliveri S, Pravettoni G. Caregiving and shared decision making in breast and prostate cancer patients: a systematic review. Current Oncology. 2023;30(1):803–823.

24. Tate A. Death and the treatment imperative: Decision-making in late-stage cancer. Soc Sci Med. 2022;306:115129.

25. Wang C, Lin C, Yang S. Hospice care improves patients’ self-decision making and reduces aggressiveness of end-of-life care for advanced cancer patients. International journal of environmental research and public health. 2022;19(23):15593.

26. Health and Retirement Study, (RAND HRS Longitudinal File 2022 (V1)) public use dataset. Produced and distributed by the University of Michigan with funding from the National Institute on Aging (grant numbers NIA U01AG009740 and NIA R01AG073289).. Accessed March 11, 2026.

27. RAND HRS Longitudinal File 2022 (V1) Produced by the RAND Center for the Study of Aging, with funding from the National Institute on Aging and the Social Security Administration.. Accessed March 11, 2026.

28. Juster FT, Suzman R. An overview of the Health and Retirement Study. J Hum Resour. 1995:S7–S56.

29. Fisher GG, Ryan LH. Overview of the health and retirement study and introduction to the special issue. Work, aging and retirement. 2018;4(1):1–9.

30. Fetherstonhaugh D, McAuliffe L, Shanley C, Bauer M, Beattie E. “Did I make the right decision?”: The difficult and unpredictable journey of being a surrogate decision maker for a person living with dementia. Dementia. 2019;18(5):1601–1614.

31. Geddis-Regan A, Errington L, Abley C, Wassall R, Exley C, Thomson R. Enhancing shared and surrogate decision making for people living with dementia: a systematic review of the effectiveness of interventions. Health Expectations. 2021;24(1):19–32.

32. Sharma RK, Dzeng E, O’Brien JM, et al. Care Decisions in the Hospital: Challenges for Family Members of Hospitalized Persons With Dementia. J Pain Symptom Manage. 2025;70(1):1–9. e3.

33. Triandafilidis Z, Carr S, Davis D, et al. Improving end-of-life care for people with dementia: a mixed-methods study. BMC palliative care. 2024;23(1):30.

34. Gupta E, Patel P. Palliative care in dementia. Annals of palliative medicine. 2024;13(4):791–807.

35. Au S, Perepeluk P, Soo A, Simon J. Defining and Characterizing Inappropriate Goals of Care Designation: A 10-year Retrospective Multicenter ICU Cohort Study. J Pain Symptom Manage. 2025;70(2):170–181. e1.

36. Birtwistle J, Russell AM, Relton SD, Easdown H, Grieve U, Allsop M. Patient and public perspectives on the availability of their health and advance care planning information to support care at the end of life: a mixed-methods questionnaire study. BMJ open. 2026;16(1):e092353.

37. Haverals R, Anthierens S, Steele Gray C, Pype P, Van den Broeck K, Boeckxstaens P. Putting “what matters to you?” into practice: a focused team-based ethnographic study on goal-oriented care. BMC Primary Care. 2026;27(1):47.

38. Yu J, Jiang L, Chen Q, et al. Conceptual Development of a Patient-Centered Consultation Framework for Multimorbidity Management in Primary Care (CARESTEP). Journal of Primary Care & Community Health. 2025;16:21501319251396213.

39. Huang H. Shared Decision-Making with Palliative Care Specialist. Journal of Hospice and Palliative Care. 2025;28(4):131.

40. Halamová L, Locock L, Maclaren A, Makin S, Phillips L. Involvement of older adults in shared decision-making on care transitions in the UK: an interpretative qualitative systematic review. Ageing & Society. 2025:1–26.

41. Taylor JO, Child CE, Sharma RK, Asirot MG, Miller LM, Turner AM. Supportive care decision-making processes of persons with dementia and their caregivers. Dementia. 2023;22(8):1695–1717.

42. Ernecoff NC, Yun HY, McCreedy E, Hanson LC, Mitchell SL. A pragmatic approach to identifying goal-concordant care for nursing home residents with Alzheimer’s disease or related dementias. Journal of the American Medical Directors Association. 2024;25(11).

43. Jacobson E, O’Callaghan T, Tian S, et al. Assessing the quality and impact of goals of care documentation before intensive care unit transfer. J Pain Symptom Manage. 2025.

44. Daly J, Schmidt M, Thoma K, Xu Y, Levy B. How well are serious illness conversations documented and what are patient and physician perceptions of these conversations? J Palliat Care. 2022;37(3):332–340.

45. van der Steen JT, Van den Block L, Nakanishi M, et al. Optimizing advance care planning in dementia: recommendations from a 33-country Delphi study. J Pain Symptom Manage. 2025;69(6):e755–e772.

46. Ashana DC, D’Arcangelo N, Gazarian PK, et al. “Don’t talk to them about goals of care”: understanding disparities in advance care planning. The Journals of Gerontology: Series A. 2022;77(2):339–346.

47. Jones T, Luth EA, Lin S, Brody AA. Advance care planning, palliative care, and end-of-life care interventions for racial and ethnic underrepresented groups: a systematic review. J Pain Symptom Manage. 2021;62(3):e248–e260.

48. Portanova J, Ailshire J, Perez C, Rahman A, Enguidanos S. Ethnic differences in advance directive completion and care preferences: what has changed in a decade? J Am Geriatr Soc. 2017;65(6):1352–1357.

49. Estrada LV, Cohen CC, Shang J, Stone PW. Community-based advance care planning interventions for minority older adults: A systematic review. J Gerontol Nurs. 2021;47(5):26–36.

50. Bazargan M, Cobb S, Assari S. Completion of advance directives among African Americans and Whites adults. Patient Educ Couns. 2021;104(11):2763–2771.

